# COVID-19 symptoms and duration of direct antigen test positivity at a community testing and surveillance site, January 2021-2022

**DOI:** 10.1101/2022.05.19.22274968

**Authors:** Carina Marquez, Andrew D. Kerkhoff, John Schrom, Susana Rojas, Douglas Black, Anthea Mitchell, Chung-Yu Wang, Genay Pilarowski, Salustiano Ribeiro, Diane Jones, Joselin Payan, Simone Manganelli, Susy Rojas, Jonathan Lemus, Vivek Jain, Gabriel Chamie, Valerie Tulier-Laiwa, Maya Petersen, Joseph DeRisi, Diane V. Havlir

**Author notes:** **Corresponding author:** Carina Marquez, MD, MPH, Associate Professor of Medicine, Division of HIV, Infectious Diseases Global Medicine, University of California, San Francisco, Zuckerberg San Francisco General Hospital, 995 Potrero Ave, San Francisco, CA 94110. Authors contributed equally (Joseph DeRisi, Diane V. Havlir).

## Abstract

**Importance:** Characterizing clinical symptoms and evolution of community-based SARS Co-V-2 infections can inform health practitioners and public health officials in a rapidly changing landscape of population immunity and viral variants.

**Objective:** To characterize COVID-19 symptoms during the Omicron period compared to pre-Delta and Delta variant periods and assess the duration of COVID-19 BinaxNOW rapid antigen test positivity during the Omicron variant surge.

**Design, Setting, and Participants:** This public health surveillance study was undertaken between January 2021-January 2022, at a walk-up community COVID-19 testing site in San Francisco, California. Testing with BinaxNOW rapid antigen tests was available regardless of age, vaccine status, or symptoms throughout.

**Main Outcomes and Measures:** We characterized the prevalence of specific symptoms for people with a positive BinaxNOW test during the Omicron period and compared it to the pre-Delta and Delta periods. During the Omicron period, we examined differences in symptoms by age and vaccine status. Among people returning for repeat testing during Omicron period, we estimated the proportion with a positive BinaxNOW antigen test between 4-14 days from symptom onset or since first positive test if asymptomatic.

**Results:** Of 63,277 persons tested, 18,301 (30%) reported symptoms and 4,568 (25%) tested positive for COVID-19. During the Omicron period, 41.6% (3032/7283) of symptomatic testers tested positive, and the proportion reporting cough (67.4%) and sore throat (43.4%) was higher than during Delta and pre-Delta periods. Congestion was higher during Omicron (38.8%) than during the pre-Delta period and loss of taste/smell (5.3%) and fever (30.4%) were less common. Fevers and myalgias were less common among persons who had received boosters compared to unvaccinated people or those who received the primary series. Five days after symptom onset, 31.1% of people with COVID-19 stated their symptoms were similar or worsening. An estimated 80.2% of symptomatic re-testers remained positive five days after symptom onset and 60.5% after ten days.

**Conclusions and Relevance:** COVID-19 upper respiratory tract symptoms were more commonly reported during the Omicron period compared to pre-Delta and Delta periods, with differences by vaccination status. Antigen test positivity remained high after 5 days, supporting guidelines requiring a negative test to shorten the isolation period.

**Key points:** *Question:* During the Omicron period, are there differences in COVID-19 symptomatology compared to the pre-Delta and Delta periods and how long do rapid antigen tests remain positive?

*Findings:* In this community-based surveillance study we detected differences in symptomatology between the Omicron period and prior variant-periods, and by age and vaccination status. Five days after symptom onset, 80% remained positive with a BinaxNOW test.

*Meaning:* During the Omicron period, differences in symptomatology may be due to rising population immunity and a new variant. BinaxNOW positivity remained high among re-testers, which supports guidelines that use rapid tests to shorten the isolation period.

## Introduction

COVID-19 symptoms are an important entry point into testing, treatment, and isolation. Prompt entry into this sequence of care is critical to breaking chains of transmission and starting anti-viral treatment early in infection^1,2^. As population immunity and viral variants evolve in the ongoing SARS CoV-2 pandemic, understanding symptomatology, symptom duration, and the use of changing diagnostic modalities, such as rapid antigen tests, can help inform medical providers and public health leaders on clinical management and key policy questions. National studies from the United Kingdom (UK) have demonstrated changes in COVID-19 symptom profile by viral variant.^3–5^ However, there has not been a large study of symptom variation by variant wave among outpatients outside of the UK. Further, there have been relatively few data during the Omicron surge on COVID-19 symptoms, their duration, variation in adult and pediatric populations, and variation by vaccination status.

Understanding the duration of symptoms and infectiousness can inform many policy questions, including return to work guidelines, and individual decisions during COVID-19 surges. Although the US Centers for Disease Control (CDC) recommended return to work after 5 days if symptoms improve^6^, a recent study from Alaska and two studies of health care workers showed that over 50-80% of people remain rapid antigen test positive 5 days after symptom onset. These data raise concerns about persistent infectiousness after 5 days, as antigen test positivity strongly correlates with the presence of viable virus.^7–11^

Among persons testing at a walk-up COVID-19 testing site in San Francisco that serves Latinx residents^12^ – a community disproportionately affected by COVID-19^12^ – our objective was to first determine the prevalence and characteristics of specific symptoms among symptomatic COVID-19 positive persons during the Omicron period compared to the Delta and pre-Delta periods as population immunity and variants evolved. During the Omicron period we further sought to assess how symptoms varied by vaccination status and age and to characterize symptom duration and persistence of a positive BinaxNOW rapid antigen test.

## Methods

### Study setting and participants

This public health surveillance study was undertaken between January 10, 2021, and January 31, 2022 at the Unidos en Salud (UeS) neighborhood testing and vaccine site, which is located in the Mission District of San Francisco. The UeS neighborhood testing and vaccination site was co-designed through a community-academic partnership between the San Francisco Latino Task Force-Response to COVID-19 (LTF), University of California, San Francisco (UCSF), and the Chan Zuckerberg Biohub.^2,13^ The site serves predominantly low-income Latinx persons, a large majority of whom are low-wage earning frontline and essential workers.^1^ It is conveniently located near a busy transportation hub and offers free walk-up testing with no request for residency or health insurance information. Throughout the study period, we conducted community outreach consistently, messaging the importance of regular testing regardless of symptoms and vaccination status due to the elevated risk of acquiring COVID-19 among the population served by the site.

### Ethics statement

The study was conducted under a public health surveillance program that was reviewed by the UCSF Committee on Human Research and determined to be exempt and waived from IRB oversight. All participants provided informed consent in their preferred language prior to survey administration and COVID-19 testing.

### Procedures and samples

All participants or their caretakers completed a structured electronic survey capturing socio-demographics, vaccination status, symptoms (including date-of-onset and trajectory [e.g., improving, unchanged, worsening]). Trained laboratory assistants performed a bilateral anterior nasal swab for COVID-19 testing using the BinaxNOW COVID-19 Ag Card (Abbott Diagnostics) and a bilateral nasal swab for sequencing. ^14,15^ We performed genotyping of all isolates as previously described.^16^ Full genome sequences are available through GISAID and previously described for the pre-Delta period (majority being Epsilon^17^ and Alpha variants) and for the Omicron period.^16^ During the Omicron period, community health workers reminded people testing positive to repeat their test 5 days after symptom onset or their initial test to assess candidacy for shortening isolation per California Department of Public Health guidelines.^18^ Clients received a text message reminding them of this option.

### Definitions and Statistical Analysis

For analysis purposes, three time periods were defined, corresponding to distinct COVID-19 waves: (1) <u>Pre-Delta period</u>: January 10, 2021 to May 31, 2021; (2) <u>Delta period</u>: June 1, 2021 to November 30, 2021; and, (3) <u>Omicron period</u>: December 1, 2021 to January 30, 2022. Two different analysis populations were defined.

First, among symptomatic testers (n=18,301) we calculated the proportion with a positive BinaxNOW rapid antigen test and the prevalence of specific symptoms during each variant period (Omicron, Delta, and pre-Delta), stratified by rapid antigen test result. Among persons with a positive rapid antigen test we evaluated whether symptom prevalence differed between the Omicron compared to the Delta and pre-Delta periods. Finally, during the Omicron period, we also assessed symptom prevalence by age and vaccination status, and determined whether self-reported symptoms improved, worsened, or remained unchanged over days following the test.

The second analysis (n=942) sought to understand the proportion of participants during the Omicron wave with a positive repeat COVID-19 BinaxNOW rapid antigen test result. This analysis was restricted to ‘re-testers’, defined as participants who had a positive BinaxNOW rapid antigen test result on or after January 1, 2022, and had at least one additional BinaxNOW rapid antigen result two or more days after their initial positive test. For each day from 4 to 14 following symptom onset, or day since initial positive test if asymptomatic, we estimated the proportion of persons who remained positive by day. Participants whose second test was positive were assumed to be positive each day between the positive tests. Test positivity on days between a positive and a negative test was imputed in three different ways: (1) assuming a linearly decreasing probability of testing positive between their two tests (main analysis), (2) assuming that tests would have converted to negative the day after their initial positive test (lower bound of sensitivity analysis), (3) assuming that tests would have remained positive until the day before their repeat negative test (upper bound of sensitivity analysis). We further stratified analyses test by symptom and vaccine status. For completeness, we also report positivity over time among repeat testers without imputation of results between tests.

For both analysis populations, Fisher’s exact or chi-squared tests were used as appropriate to compare proportions. Kruskal-Wallis tests were used to compare medians. We report both unadjusted p-values and, for multiple comparisons of symptom prevalence across periods, vaccination, and age strata, p-values adjusted using the Benjamin-Hochberg method to control false discovery rate at 5%.

## Results

Between January 10, 2021, and January 31, 2022, 63,277 persons underwent COVID-19 testing at the UeS Neighborhood site. Overall, 18,301 (29.9%) participants across all waves reported at least one current symptom at the time of COVID-19 testing; 37.1% (n=7,823) during Omicron, 28.3% during Delta (n=5,485), and 22.8% (n=5,533) during the pre-Delta period. Test positivity was 41.6%, 8.5%, and 19.2%, respectively. Overall, the median age of testers was 32 (IQR: 21-44) years old, 12.0% were <12, 52.0% were women, and 68.5% were Latinx. During the Omicron period, the median age was 34.2, 52.6% were female, 74.3% were Latinx, 73.0% of reported a household income of <$50,000 per year. We detected small differences in who sought testing by variant-period according to age, gender, and race (Supplementary Table 1). The proportion of unvaccinated participants increased throughout variant periods, during Omicron 12.7% of symptomatic testers were unvaccinated compared to 39.7% during the Delta period, and 93.3% during the pre-Delta period, p<0.01 (Supplementary Table 1).

### Symptom Profile of People Testing Positive for COVID-19

During the Omicron period (n=3,032), the most common symptoms reported by persons testing positive for COVID-19 by BinaxNOW rapid antigen tests were cough (67.4%), sore throat (43.4%), congestion (38.8%), headache (35.5%), while loss of smell/taste (5.3%), nausea (5.0%) and diarrhea (4.8%) were the least commonly reported (Table 1A). The proportion of symptomatic COVID-19 positive persons reporting cough and sore throat was higher during the Omicron period relative to Delta and pre-Delta periods, while fever, and loss of taste/smell decreased relative to Delta and pre-Delta periods (Table 1A, Supplementary Figure 1, p<0.05 for all comparisons). Congestion was more common among symptomatic people testing positive during the Omicron period compared to pre-Delta (p<0.001). The leading COVID-19 symptoms among symptomatic people testing positive during the Omicron period (cough, sore throat, and congestion) were also common among symptomatic people who tested negative (Table 1, Supplementary Figure 1).

**Table 1:** Symptoms reported among symptomatic people testing positive and negative with the BinaxNOW rapid antigen test by, A. Variant period: Pre-Delta (January 10-May 31, 2021), Delta (June 1, 2021-November 30, 2021), Omicron Period.(December 1, 2021-January 30,20222)

|  | Overall<br>(N=18,301) | Pre-Delta<br>(N=5,533) |  | Delta<br>(N=5,485) |  | Omicron<br>(N=7,283) |  | Omicron vs.<br>pre-Delta<br>among<br>positives | Omicron vs.<br>Delta among<br>positives |
| --- | --- | --- | --- | --- | --- | --- | --- | --- | --- |
|  |  | Positive<br>(n=1065) | Negative<br>(n=4468) | Positive<br>(n=468) | Negative<br>(n=5017) | Positive<br>(n=3032) | Negative<br>(n=4251) | P-value<br>(B-H<br>corrected) | P-value<br>(B-H<br>corrected) |
| **Symptoms** |  |  |  |  |  |  |  |  |  |
| Fever | 3527 (19.3) | 369 (34.7) | 672 (15.0) | 172 (36.8) | 780 (15.6) | 921 (30.4) | 613 (14.4) | 0.010<br>(0.02) | 0.006<br>(0.015) |
| Cough | 8885 (48.6) | 546 (51.3) | 1507 (33.7) | 281 (60.0) | 2356 (47.0) | 2044 (67.4) | 2151 (50.6) | <0.001<br>(0.003) | 0.002<br>(0.006) |
| Shortness of breath | 1386 (7.6) | 93 (8.7) | 409 (9.2) | 42 (9.0) | 346 (6.9) | 241 (8.0) | 255 (6.0) | 0.42<br>(0.51) | 0.45<br>(0.52) |
| Fatigue | 3404 (18.6) | 176 (16.5) | 926 (20.7) | 107 (22.9) | 879 (17.5) | 594 (19.6) | 722 (17.0) | 0.028<br>(0.051) | 1<br>(1) |
| Myalgia | 3896 (21.3) | 322 (30.2) | 978 (21.9) | 141 (30.1) | 829 (16.5) | 868 (28.6) | 758 (17.8) | 0.32<br>(0.41) | 0.51<br>(0.56) |
| Headache | 6064 (33.1) | 437 (41.0) | 1693 (37.9) | 167 (35.7) | 1420 (28.3) | 1075 (35.5) | 1272 (29.9) | 0.001<br>(0.003) | 0.92<br>(0.96) |
| Loss of taste/smell | 951 (5.2) | 183 (17.2) | 207 (4.6) | 96 (20.5) | 177 (3.5) | 160 (5.3) | 128 (3.0) | <0.001<br>(0.003) | <0.001<br>(0.003) |
| Sore throat | 6570 (35.9) | 315 (29.6) | 1399 (31.3) | 136 (29.1) | 1821 (36.3) | 1316 (43.4) | 1583 (37.2) | <0.001<br>(0.003) | <0.001<br>(0.003) |
| Congestion | 6217 (34.0) | 294 (27.6) | 1216 (27.2) | 193 (41.2) | 1866 (37.2) | 1177 (38.8) | 1471 (34.6) | <0.001<br>(0.003) | 0.32<br>(0.41) |
| Nausea | 1108 (6.1) | 75 (7.0) | 295 (6.6) | 29 (6.2) | 324 (6.5) | 150 (5.0) | 235 (5.5) | 0.010<br>(0.020) | 0.25<br>(0.37) |
| Diarrhea | 1192 (6.5) | 65 (6.1) | 425 (9.5) | 28 (6.0) | 304 (6.1) | 144 (4.8) | 226 (5.3) | 0.08<br>(0.135) | 0.25<br>(0.37) |

Abbreviations: B-H corrected= Benjamin-Hochberg method

During the Omicron period, 47.7% of symptomatic children (<12 years) with COVID-19 reported only one symptom. Reporting a single symptom was less common among adults (37.7%, p<0.001) and adolescents (31.8%, p<0.001) (Supplemental Table 2). Among children with COVID-19, cough, fever, sore throat, and congestion were the most common symptoms. Loss of taste/smell was uncommon among children (0.3%) compared to adolescents (5.8%, p<0.001) and adults (5.8%, p<0.001) (Table 1B).

We also found differences in symptoms according to vaccination status (Table 3). Among boosted persons, congestion was more common, and fever was less common compared to both unvaccinated people and vaccinated/non-boosted people (p<0.05 for all comparisons; Table 1C); myalgias were also less common among boosted persons compared to vaccinated/non-boosted persons (p=0.01). The prevalence of other symptoms did not differ by vaccination status.

**Table 2:**
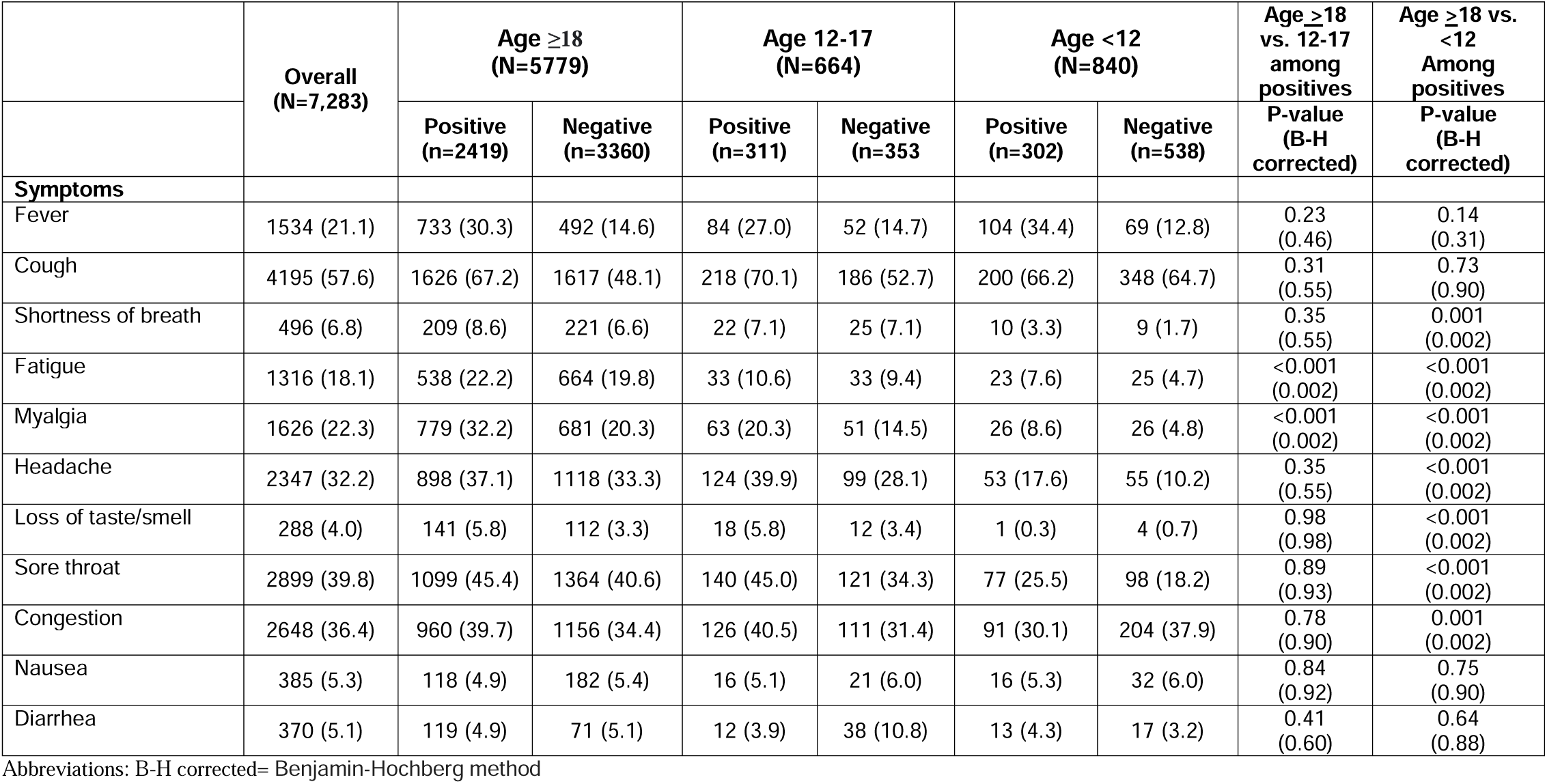
Symptoms reported among symptomatic people testing positive and negative with the BinaxNOW rapid antigen test during the Omicron period (December 1, 2021-January 30,20222), stratified by age group.

**Table 3:** Symptoms reported among symptomatic people testing positive and negative with the BinaxNOW rapid antigen test during the Omicron period (December 1, 2021-January 30,20222), stratified by vaccination status.

|  | Overall<br>(N=5424) | Unvaccinated<br>(N=233) |  | Vaccinated, not boosted<br>(N=3817) |  | Vaccinated, Boosted<br>(N=1374) |  | Boosted vs.<br>Unvaccinated<br>among<br>positives | Boosted vs.<br>Vaccinated,<br>not boosted<br>among<br>positives |
| --- | --- | --- | --- | --- | --- | --- | --- | --- | --- |
|  |  | Positive<br>(n=116) | Negative<br>(n=117) | Positive<br>(n=1705) | Negative<br>(n=2112) | Positive<br>(n=432) | Negative<br>(n=942) | P-value<br>(B-H<br>corrected) | P-value<br>(B-H<br>corrected) |
| **Symptoms** |  |  |  |  |  |  |  |  |  |
| Fever | 1162 (21.4) | 42 (36.2) | 16 (13.7) | 559 (32.8) | 343 (16.2) | 97 (22.5) | 105 (11.2) | 0.003<br>(0.01) | <0.001<br>(0.01) |
| Cough | 3060 (56.4) | 71 (61.2) | 53 (45.3) | 1191 (69.9) | 1086 (51.4) | 268 (62.0) | 391 (41.5) | 0.87<br>(1) | 0.002<br>(0.01) |
| Shortness of<br>breath | 408 (7.5) | 10 (8.6) | 7 (6.0) | 150 (8.8) | 150 (7.1) | 36 (8.3) | 55 (4.8) | 0.92<br>(1) | 0.76<br>(0.93) |
| Fatigue | 1144 (21.1) | 33 (28.5) | 25 (21.4) | 372 (21.8) | 410 (19.4) | 104 (24.1) | 200 (21.2) | 0.33<br>(0.50) | 0.31<br>(0.50) |
| Myalgia | 1383 (25.5) | 41 (35.3) | 25 (21.4) | 580 (34.0) | 440 (20.8) | 115 (26.6) | 182 (19.3) | 0.06<br>(0.19) | 0.003<br>(0.01) |
| Headache | 1921 (35.4) | 49 (42.2) | 42 (35.9) | 646 (37.9) | 719 (34.0) | 153 (35.4) | 312 (33.1) | 0.18<br>(0.40) | 0.34<br>(0.50) |
| Loss of<br>taste/smell | 240 (4.4) | 12 (10.3) | 7 (6.0) | 99 (5.8) | 76 (3.6) | 25 (5.8) | 21 (2.2) | 0.08<br>(0.22) | 0.99<br>(1) |
| Sore throat | 2325 (42.9) | 48 (41.4) | 34 (29.1) | 776 (45.5) | 850 (40.3) | 208 (48.2) | 409 (43.4) | 0.20<br>(0.40) | 0.33<br>(0.50) |
| Congestion | 2019 (37.2) | 40 (34.5) | 32 (27.4) | 668 (39.2) | 701 (33.2) | 207 (47.9) | 371 (39.4) | 0.010<br>(0.04) | 0.001<br>(0.01) |
| Nausea | 290 (5.4) | 7 (6.0) | 6 (5.1) | 89 (5.2) | 119 (5.6) | 18 (4.2) | 51 (5.4) | 0.39<br>(0.50) | 0.37<br>(0.50) |
| Diarrhea | 277 (5.1) | 10 (8.6) | 2 (1.7) | 83 (4.9) | 112 (5.3) | 21 (4.9) | 49 (5.2) | 0.12<br>(0.29) | 1<br>(1) |

Abbreviations: B-H corrected= Benjamin-Hochberg method

### Time to improvement of COVID-19 symptoms during the Omicron period

Next, we evaluated the trajectory of symptoms among persons testing COVID-19 positive during the Omicron period (**Figure 1A**; n=1,613). Among participants tested five days after symptom onset, 63.0% (95%CI: 56.6-69.2) of persons had improving symptoms by day of testing, symptoms were similar in 31.1% (95%CI: 25.3%-37.4%) and in 5.9% were worsening (95%CI: 3.3%-9.7%). Among participants testing ten days after symptom onset, symptoms were improving in 82.2% (95%CI: 56.6%-69.2%), were similar in 17.8% (95%CI: 9.8%-28.5%) and were worsening in 0% (95%CI: 0%-4.9%). Symptom trajectory was similar across vaccination status or age (**Figure 1B, 1C)**.

**Figure 1:**
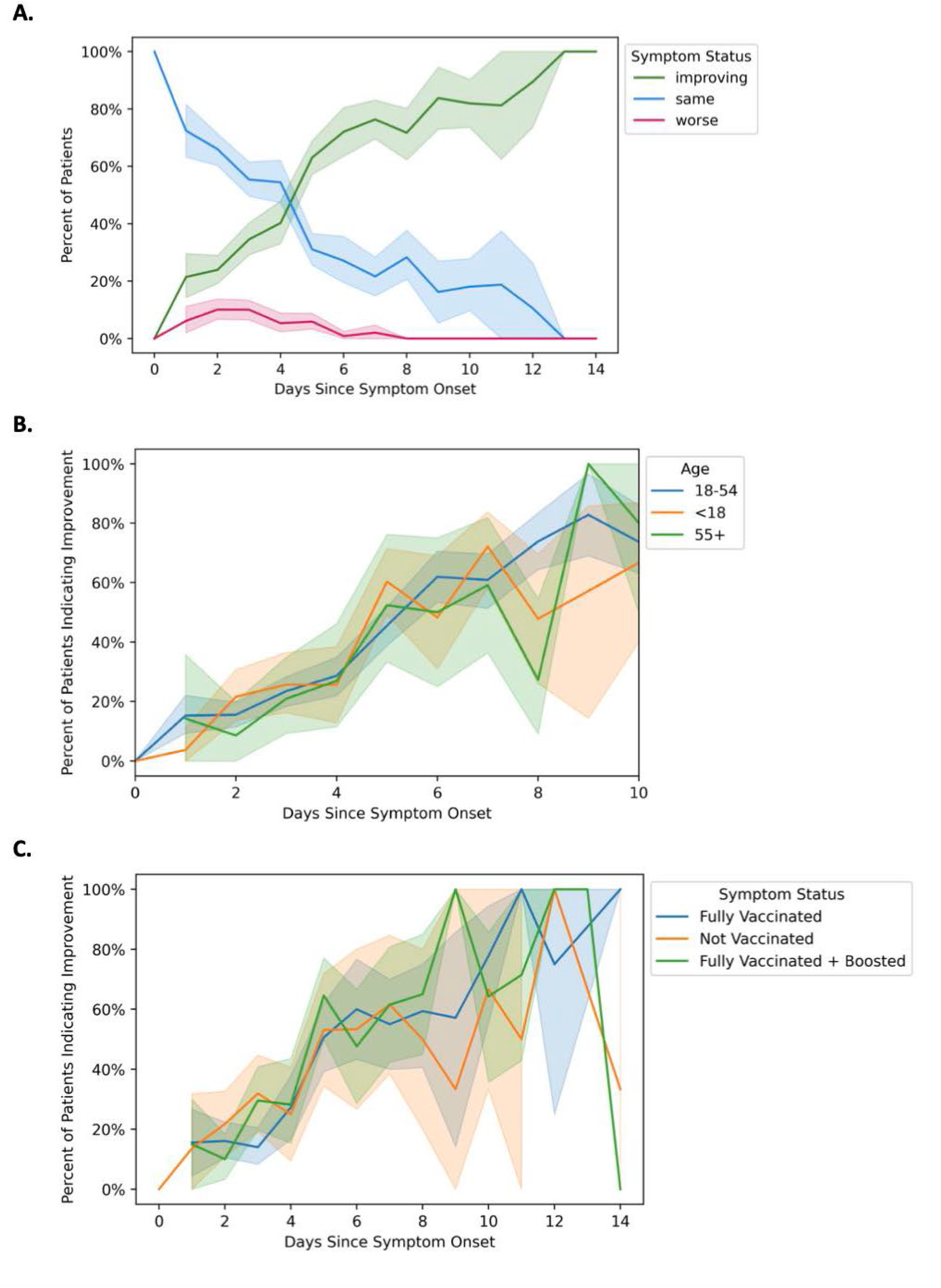
Proportion of COVID-19 positive clients during Omicron period indicating improvement, no change, or worsening of symptoms at time of test, among symptomatic BinaxNOW positive participants testing within 14 days of symptom onset, A. Overall, B. by Age Group and C. and by Vaccination Status

### COVID-19 repeat testing results

Among persons who undertook COVID-19 re-testing, an estimated 65% (95%CI: 62%-69%) people remained test-positive 5 days after symptom onset or 5 days from their initial positive test if they were asymptomatic (Figure 2A). Among symptomatic persons, an estimated 80% (95% CI: 76%-84%) and 35% (95%CI: 30%-40%) remained positive on day 5 and 10 respectively. Among persons asymptomatic at testing, an estimated 49% (95% CI: 34-55%) and 19% (95% CI, 5%-42%) remained positive on day 5 and 10. Repeat test positivity was higher among symptomatic vs. asymptomatic testers on day 5 and 10. (Figure 2B). We found no difference in trajectory of test positivity over time by vaccination status (Figure 2C). Similar results were found using non-imputed values for interim test positivity (Supplementary Figure 2A-2C).

**Figure 2:**
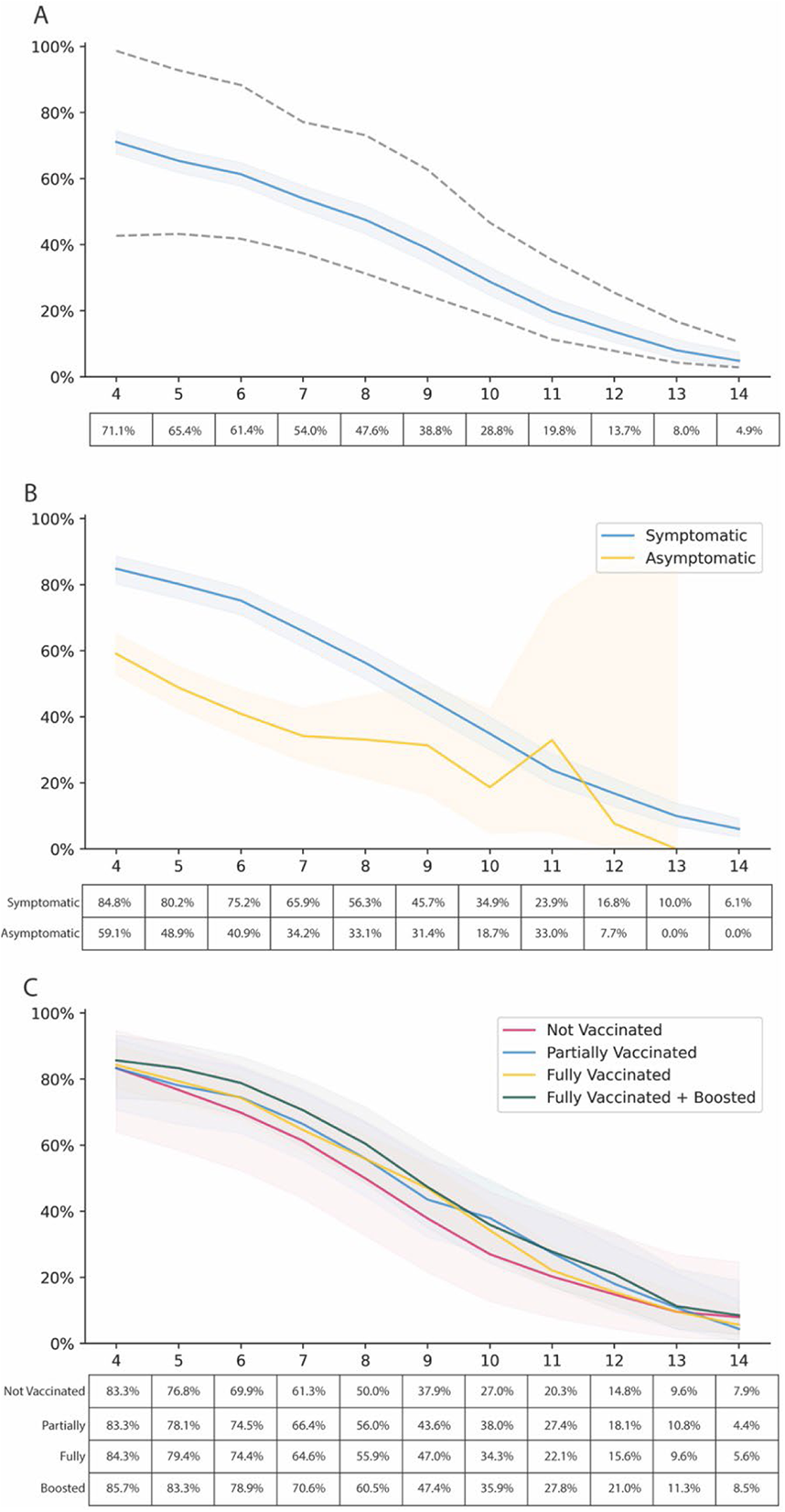
BinaxNOW positivity among 942 repeat testers with COVID-19 during the Omicron period by day of symptom onset (if symptomatic) or day since initial positive test (if asymptomatic). COVID-19 status between a participant’s tests is inferred to be positive (if second test is positive) or with a linearly decaying probability of being positive (if second test is negative). Panel A shows overall, including symptomatic and asymptomatic people with dotted lines indicating upper and lower bounds of sensitivity analysis. Sensitivity analysis assumes participants whose second test is negative either immediately became negative after their first test (lower bound) or remained positive until the day before their second test (upper bound). Panel B shows results stratified by symptom status. Panel C show results stratified by self-report vaccination status as of their first test.

## Discussion

Over the course of three different COVID-19 surges, each with a different predominant SARS-CoV-2 variant, and against a backdrop of rising population immunity, we found a shift towards predominantly upper respiratory tract symptoms with the Omicron variant among symptomatic persons presenting for COVID testing at a low-barrier, walk-up testing site. Omicron symptom trajectories were heterogenous, with a third of people reporting no improvement or worsening of symptoms when they returned for repeat testing with a BinaxNOW rapid antigen test after 5 days. During the Omicron surge, 80% of symptomatic persons with COVID-19 returning for repeat testing were BinaxNOW positive after 5 days, and 35% still positive after 10 days, regardless of vaccine status. These results provide updated clinical data on COVID-19 outpatients and rapid antigen test positivity to guide safe return-to-work guidelines.

In over 3,000 symptomatic people testing positive for COVID-19 during an Omicron surge, upper respiratory tract symptoms (sore throat, congestion) were more common than during pre-Delta and Delta periods. The loss of taste and smell - often used by clinicians to help distinguish SARS Co-V 2 from other viral illness - was much less common during the Omicron period (5.3%) compared to the pre-Delta (17.2%) and Delta (20.5%) periods. This shift in symptomatology seen from pre-Delta to Omicron is likely due to both the increase in population immunity during the Omicron surge and biologic characteristics of the Omicron variant, which in vitro-studies suggest replicates better in bronchial tissue versus deeper lung sites.^19^ During the Omicron surge, symptoms also varied by vaccine status; congestion was more common, and fever and myalgia were less common among boosted persons compared to partially or unvaccinated persons.

Our findings are concordant with a population-representative household study conducted during the Omicron period in the United Kingdom (UK) ^5^ that found a predominance of upper respiratory tract symptoms. Additionally, our data expands existing knowledge on COVID-19 symptomatology by describing differences in symptomatology by vaccine status and age-group. Assessing the ability of symptoms to predict test positivity was outside the scope of our analysis. However, similar to findings from the UK findings,^5^ we found the background rate of common COVID-19 symptoms during the Omicron period to be high among both people who tested both positive and negative. These findings emphasizing the importance of ensuring low-barrier testing is available regardless of the tester’s symptom profile.

This study is one of the few in the Omicron era to describe symptoms of mild-moderate infection in children. While symptoms were heterogenous in children, over 40% of children under the age of 12 had only one symptom, with fever, cough, and congestion being the most common. It is important to recognize that children, especially those under five, may present with only one symptom. Thus, parents and health care providers should have a low-threshold to test children with any symptom potentially concerning for COVID-19. Rapid and easily accessible testing is an important strategy to keep children in school, while minimizing transmission.^21,22^

Despite lower frequencies of constitutional symptoms during the Omicron surge, it is noteworthy that recovery varied and 31% of patients did not report perceived improvement by day 5. During the Omicron surge, frontline workers were urged to return to work to meet staffing needs in occupations such as transport, home health care, and the food service industry. During surges of the magnitude of Omicron, frontline workers can be urged to return to work to meet staffing needs in occupations such as transport, home health care, and the food service industry. Despite not having illness serious enough to merit hospitalization, many persons with Omicron still felt ill, yet felt pressure by their employers to return to work. Safeguards routinely in place in the formal sector in the United States, are often unavailable in the informal sector.^23,24^ Pandemic planning must account for these health and economic pressures in future surges.

Of persons with COVID-19 re-testing on Day 5, 80% of symptomatic people and 41% of asymptomatic people remained positive. This number is nearly identical to a community-based sample of people seeking testing in Alaska^25^ and similar to the findings in two cohorts of health care workers^26,27^. This high proportion of positive repeat tests is not surprising, given existing data on viral dynamics of Omicron and other variants. In a predominantly boosted cohort from the National Basketball Association (NBA), viral trajectories of Omicron were heterogenous, but the duration of the acute phase (proliferation and clearance) was 9.9 days and similar to prior variants, including Delta.^28,29^ Additionally, in this cohort of predominantly healthy, boosted NBA players with infections due to the Omicron variant, only 52% of people had a cycle threshold of <30 5 days after symptom onset.^28^

Interestingly, we found that COVID-19 test positivity remained high even 10 days following symptom onset, where more than 30% or people with symptomatic COVID-19 during their initial still had a positive rapid antigen test. Additional epidemiologic studies are needed to assess whether people remain infectious at this juncture. However, prior studies show a strong correlation between rapid antigen positivity and viable virus,^7–9,30^ even in the clearance phase and after 10 days of symptom onset ^9,10,31^-suggesting infectiousness beyond 10 days may be possible, though less common.

Positive rapid antigen tests correlate to having viable virus, and thus capture persons with the highest degree of infectivity to others.^7–9,11^ The CDC recommends that people can leave isolation with a well-fitting mask after 5 days of symptoms if symptoms are improving, regardless of repeat testing.^6^ If a public health goal is to break chains of SARS CoV-2 transmission, our data strongly support current California Department of Public Health guidelines^18^ that require a rapid antigen test to shorten the 10-day isolation period.

Strengths of our study include the use of data spanning multiple COVID-19 pandemic waves from one high-throughput, low-barrier, community-based testing site that predominantly serves a highly impacted low-income Latinx population. Further, symptom ascertainment and testing procedures did not change during the surveillance period. Our study had certain limitations. Although it was cross-sectional, our data provide an important synthesis of symptomatology within a diverse population of people seeking community testing. Second, symptoms and timing of onset were self-reported which, may introduce bias; however, symptom data were collected prior to persons knowing their COVID-19 test result and thus would not be expected to result in differential bias between those who were COVID-19 positive and negative. Additionally, we did not characterize the severity of COVID-19 symptoms across waves, and even a single very severe symptom can cause substantially greater morbidity in an individual than multiple mild symptoms. Finally, testing was undertaken with rapid antigen tests, and thus may have resulted in misclassification of individuals during the earliest phases of their illness. Nonetheless, strict quality control measures were implemented to ensure accuracy of results and the use of rapid antigen tests.

In summary, the clinical presentation of symptomatic persons changed over time during several COVID surges characterized by increasing population immunity and different SARS CoV-2 variants. During Omicron, when population immunity was higher, persons with symptoms often did not see improvement after 5 days and antigen tests frequently remained positive. These findings highlight the importance of work assurances to protect workers and requirements for rapid antigen testing to shorten isolation to protect the workplace. With the dynamic landscape of host immunity and viral evolution, we need real time data from diverse populations for clinicians, public health officials and the community to develop optimal strategies to mitigate the health, economic and societal effects for SARS CoV-2 and to ensure that inequities in this disease are not exacerbated.

## Data Availability

All data produced in the present work are contained in the manuscript

## Figure Captions

**Supplementary Figure 1**. Prevalence of selected symptoms among symptomatic people testing positive and negative with the BinaxNOW rapid antigen test during pre-Delta (January 10-May 31, 2021), Delta (June 1, 2021-November 30, 2021), and Omicron period (December 1, 2021-January 30, 2021).

**Supplementary Figure 2:** BinaxNOW positivity among 942 repeat testers with COVID-19 during the Omicron period. by day of symptom onset (if symptomatic) or day since initial positive test (if asymptomatic). A. Overall, includes symptomatic people and asymptomatic people. B. Stratified by symptom status. Symptomatic include people who report symptoms at the time of their initial positive test and who note a symptom onset date and asymptomatic people include people who do not report symptoms at the time of testing. C. Stratified by self-reported vaccination status.

**Supplementary Table 1:**
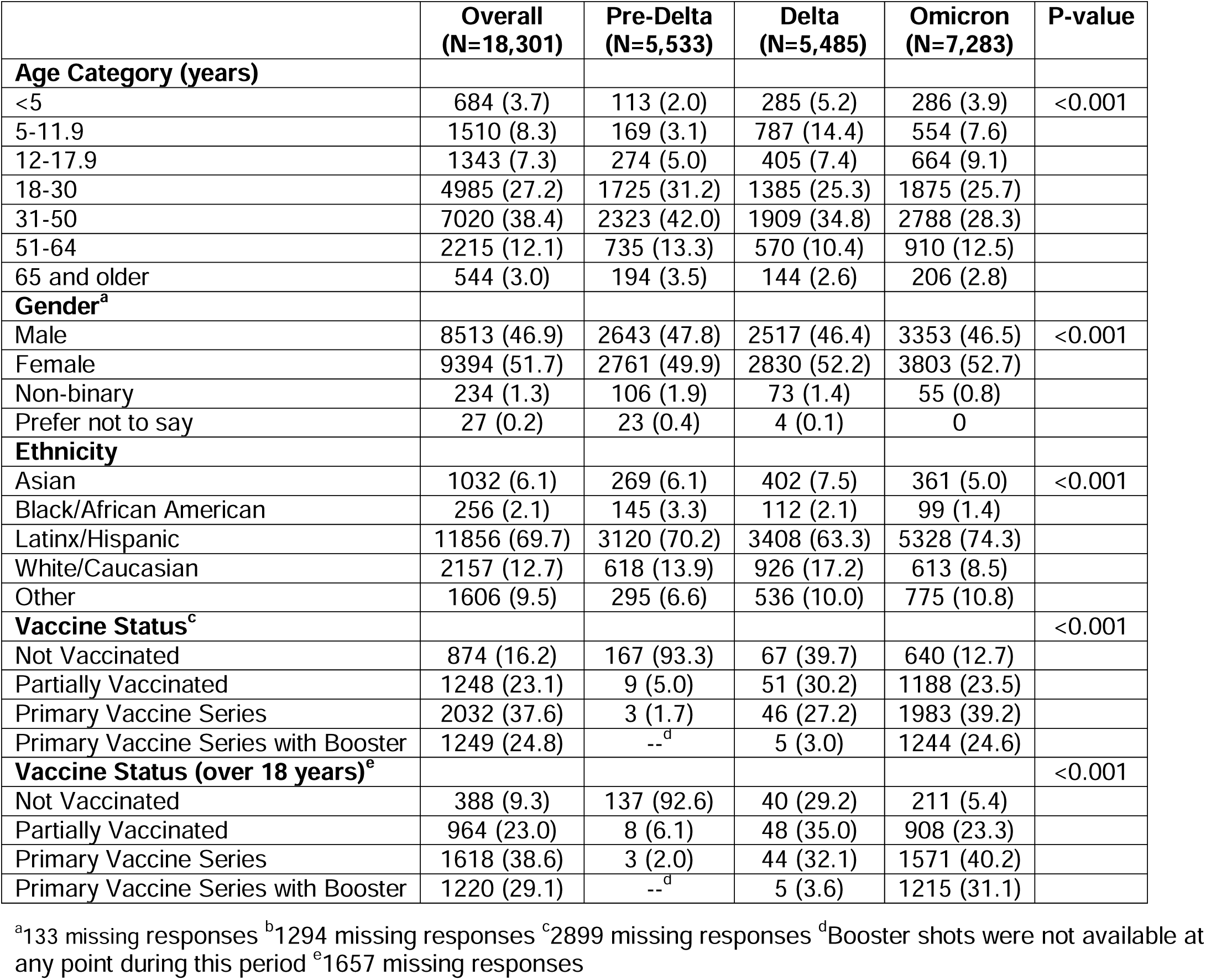
Demographics of symptomatic people seeking testing from January 2020-January 2021, by variant-period.

**Supplementary Table 2.** Proportion symptomatic participants reporting only one of the following symptoms the time of testing during the Omicron period ((December 1, 2021-January 30, 2021)

| Symptom Type | Overall<br>(N=3032) | Age ≥18<br>(N=2419) | Age 12-17<br>(N=311) | Age <12<br>(N=302) | P-value |
| --- | --- | --- | --- | --- | --- |
| Fever | 74 (2.4) | 45 (1.9) | 3 (1.0) | 26 (8.6) | <0.001 |
| Cough | 477 (15.7) | 346 (14.3) | 48 (15.4) | 83 (27.5) | 0.030 |
| Shortness of breath | 4 (0.1) | 4 (0.2) | 0 | 0 | 1 |
| Fatigue | 12 (0.4) | 10 (0.4) | 0 | 2 (0.7) | 0.41 |
| Myalgia | 25 (0.8) | 22 (0.9) | 2 (0.6) | 1 (0.3) | 0.76 |
| Headache | 51 (1.7) | 37 (1.5) | 11 (3.5) | 3 (1.0) | 0.036 |
| Loss of taste/smell | 5 (0.2) | 4 (0.2) | 1 (0.3) | 0 | 0.68 |
| Sore throat | 168 (5.5) | 147 (6.1) | 17 (5.5) | 4 (1.3) | 0.001 |
| Congestion | 140 (4.6) | 100 (4.1) | 17 (5.5) | 23 (7.6) | 0.019 |
| Nausea | 4 (0.1) | 3 (0.1) | 0 | 1 (0.3) | 0.39 |
| Diarrhea | 2 (0.1) | 1 (0.0) | 0 | 1 (0.3) | 0.20 |
| ***Any of the above symptoms***<br>***in isolation*** | 962 (31.7) | 719 (29.7) | 99 (31.8) | 144 (47.7) | <0.001 |

