## Supplementary Figure 1 for "COVID-19 symptoms and duration of direct antigen test positivity at a community testing and surveillance site, January 2021-2022"

**Supplementary Figure 1:** Prevalence of selected symptoms among symptomatic people testing positive and negative with the BinaxNOW rapid antigen test during pre-Delta (January 10-May 31, 2021), Delta (June 1, 2021- November 30, 2021), and Omicron period (December 1, 2021-January 30, 2021).

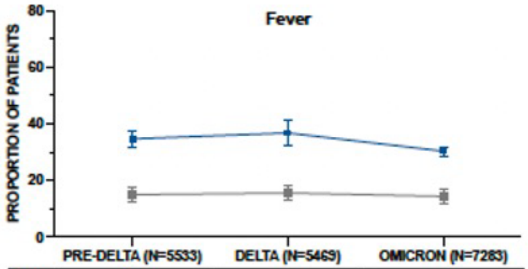


| | | | |
|---|---|---|---|
| Pos. | 34.7% | 36.8% | 30.4% |
| Neg. | 15.0% | 15.6% | 14.4% |

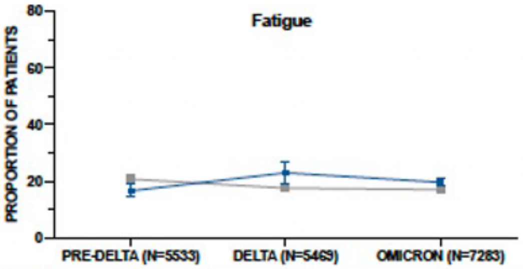


| | | | |
|---|---|---|---|
| Pos. | 16.5% | 22.9% | 19.6% |
| Neg. | 20.7% | 17.5% | 17.0% |

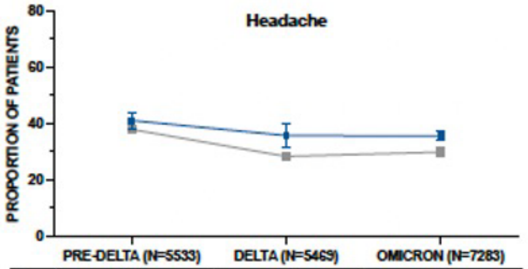


| | | | |
|---|---|---|---|
| Pos. | 41.0% | 35.7% | 35.5% |
| Neg. | 37.9% | 28.3% | 29.9% |

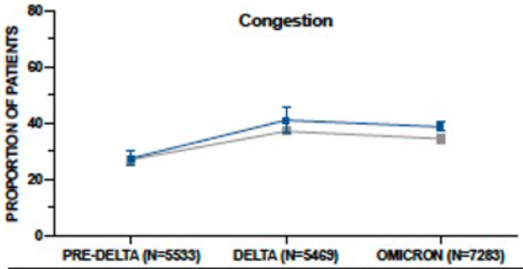


| | | | |
|---|---|---|---|
| Pos. | 27.6% | 41.2% | 38.8% |
| Neg. | 27.2% | 37.2% | 34.6% |

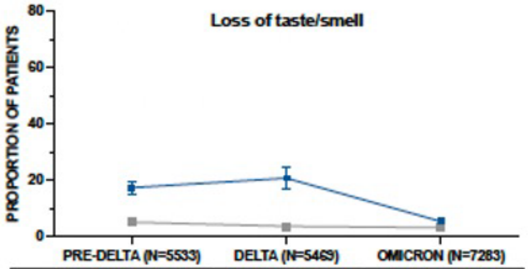


| | | | |
|---|---|---|---|
| Pos. | 17.2% | 20.5% | 5.2% |
| Neg. | 5.6% | 3.5% | 3.0% |

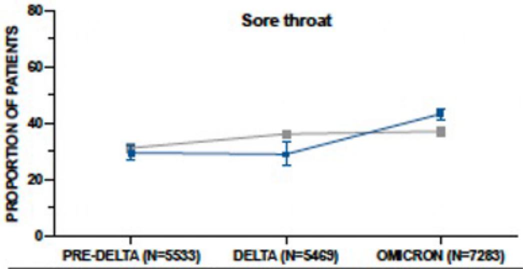


| | | | |
|---|---|---|---|
| Pos. | 29.6% | 29.1% | 43.4% |
| Neg. | 31.3% | 36.3% | 37.2% |

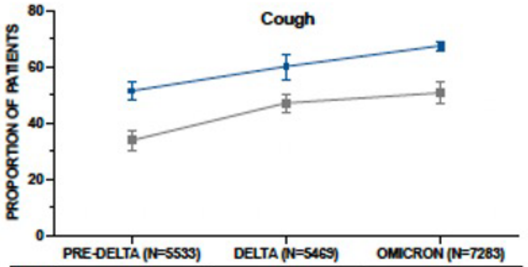


| | | | |
|---|---|---|---|
| Pos. | 51.3% | 60.0% | 30.4% |
| Neg. | 33.7% | 47.0% | 50.6% |

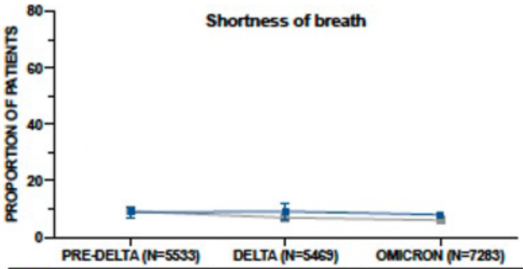


| | | | |
|---|---|---|---|
| Pos. | 8.7% | 9.0% | 7.9% |
| Neg. | 9.2% | 6.9% | 6.0% |

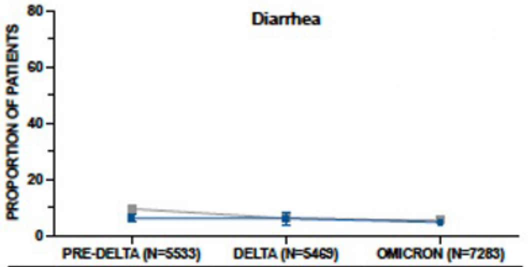


| | | | |
|---|---|---|---|
| Pos. | 6.1% | 6.0% | 4.7% |
| Neg. | 9.5% | 6.1% | 5.3% |

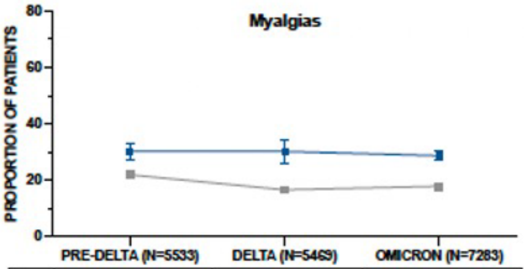


| | | | |
|---|---|---|---|
| Pos. | 30.2% | 30.1% | 28.6% |
| Neg. | 21.9% | 16.5% | 17.9% |
