## Supplementary Figure 2 for "COVID-19 symptoms and duration of direct antigen test positivity at a community testing and surveillance site, January 2021-2022"

**Supplementary Figure 2:** BinaxNOW positivity among 942 repeat testers with COVID-19 during the Omicron period. by day of symptom onset (if symptomatic) or day since initial positive test (if asymptomatic). A. Overall, includes symptomatic people and asymptomatic people. B. Stratified by symptom status. Symptomatic include people who report symptoms at the time of their initial positive test and who note a symptom onset date and asymptomatic people include people who do not report symptoms at the time of testing. C. Stratified by self-reported vaccination status.

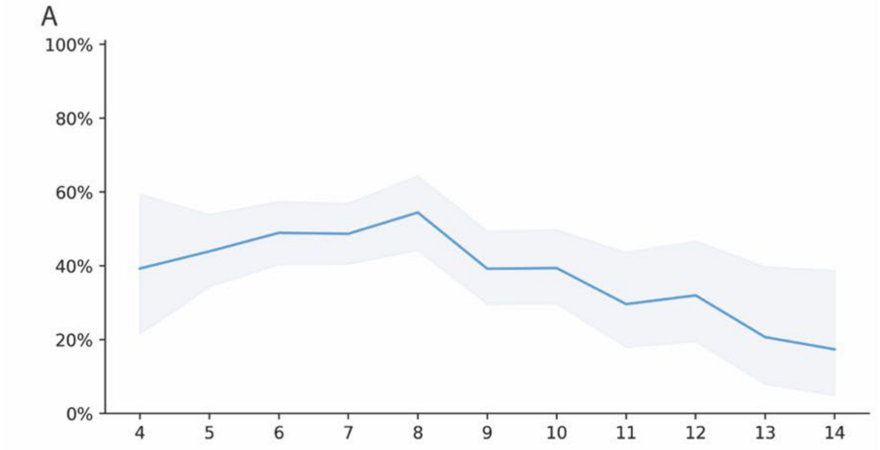


| | | | | | | | | | | |
|---|---|---|---|---|---|---|---|---|---|---|
| 39.3% | 43.5% | 48.9% | 48.7% | 54.5% | 39.2% | 39.4% | 29.6% | 32.0% | 20.7% | 17.4% |

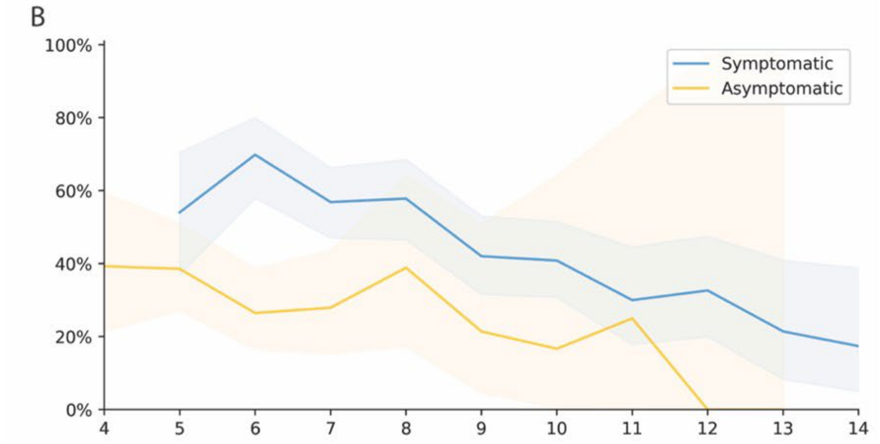


| | | | | | | | | | | | |
|---|---|---|---|---|---|---|---|---|---|---|---|
| Symptomatic | -- | 54.1% | 69.9% | 56.9% | 57.8% | 42.0% | 40.9% | 30.0% | 32.7% | 21.4% | 17.4% |
| Asymptomatic | 39.3% | 38.6% | 26.5% | 27.9% | 38.9% | 21.4% | 16.7% | 25.0% | 0% | 0% | -- |

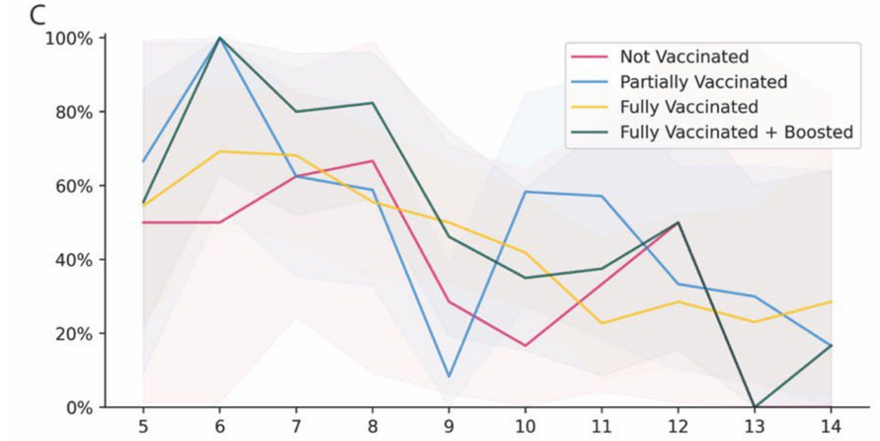


| | | | | | | | | | | |
|---|---|---|---|---|---|---|---|---|---|---|
| Not Vaccinated | 50.0% | 50.0% | 62.5% | 66.7% | 28.6% | 16.7% | 33.3% | 50.0% | 0.0% | 0.0% |
| Partially | 66.7% | 100% | 62.5% | 58.8% | 8.3% | 58.3% | 57.1% | 33.3% | 30.0% | 16.7% |
| Fully | 54.5% | 69.2% | 68.2% | 55.6% | 50.0% | 41.9% | 22.7% | 28.6% | 23.1% | 28.6% |
| Boosted | 55.6% | 100% | 80.0% | 82.4% | 46.1% | 35.0% | 37.5% | 50.0% | 0.0% | 16.7% |
